# Lung cancer mortality in the wake of the changing smoking epidemic: a descriptive study of the global burden in 2020 and 2040

**DOI:** 10.1101/2022.12.29.22284032

**Authors:** András Wéber, Eileen Morgan, Jerome Vignat, Mathieu Laversanne, Margherita Pizzato, Harriet Rumgay, Deependra Singh, Péter Nagy, István Kenessey, Isabelle Soerjomataram, Freddie Bray

## Abstract

**Objectives:** Lung cancer is the leading cause of cancer death in 2020, responsible for almost one in five (18.0%) deaths. This paper provides an overview of the descriptive epidemiology of lung cancer on the basis of national mortality estimates for 2020 from the International Agency for Research on Cancer (IARC), and in the context of recent tobacco control policies.

**Methods:** Age-standardized mortality rates per 100,000 person-years of lung cancer for 185 countries by sex were obtained from the GLOBOCAN 2020 database and stratified by Human Development Index (HDI). Lung cancer deaths were projected to 2040 based on demographic changes alongside scenarios of annually increasing, stable or decreasing rates from the baseline year of 2020.

**Results:** Lung cancer mortality rates exhibited marked variations by geography and sex. Low HDI countries, many of them within sub-Saharan Africa, tend to have low levels of mortality and an upward trend in lung cancer deaths is predicted for both sexes until 2040 according to demographic projections, irrespective of trends in rates. In very high HDI countries, including Europe, Northern America and Australia/New Zealand, there are broadly decreasing trends in men whereas in women, rates are still increasing or reaching a plateau.

**Conclusion:** The current and future burden of lung cancer in a country or region largely depends on the present trajectory of the smoking epidemic in its constituent populations, with distinct gender differences in smoking patterns, both in transitioning and transitioned countries. Further elevations in lung cancer mortality are expected worldwide, raising important social and political questions, especially in low- and middle-income countries.

**Strengths and limitations of this study:** 

**Strengths:** 

**This study:**

- presents a detailed profile of the present LC burden in men and women worldwide according to national levels of human development.
- applies a simple projection to estimate the future lung cancer mortality burden in 2040.
- discusses the results in the context of key risk factors for lung cancer, particularly the continually evolving smoking epidemic.

**Limitations:** 

**This study:**

- is hampered by the limited availability of local cause of death information from national vital registration sources, particularly in transitioning countries.

## Introduction

Lung cancer (LC) ranks as the most frequent form of cancer death and premature cancer death (ages 30-69) with a uniformly low 5-year survival, even in high-income countries [1]. With one-fifth of the present cancer mortality worldwide due to LC – an estimated 1.8 million deaths in 2020 [2] – the key determinant remains tobacco consumption. Up to 9 in 10 LC cases are caused by smoking in high-income settings, while mortality increases with number of cigarettes smoked and smoking duration [3]. Lopez at al. drew attention to the phases of the global smoking epidemic and the subsequent impact of smoking on LC occurrence by sex [4]; men and women remain in very different phases of the smoking epidemic, as reflected in disease rates by birth cohort. Recent reports have generally described marked variations in rates between sexes, with stable or decreasing rates found predominantly among male while increasing rates among female populations [5,6].

An emerging pattern is a higher rate of LC incidence among young females than males across geographic areas and income levels, that is not fully explained by sex-specific differences in smoking prevalence [7]. Such temporal patterns forewarn of a higher LC burden in women than men at older ages in the decades to follow, especially in higherlincome settings. Women have been increasingly targeted in marketing campaigns, particularly in transitioning countries, while social constraints that precluded women taking up the habit are weakening [8]; still, smoking prevalence among women varies markedly, with, for example, a small proportion of women in China current smokers, in absolute terms and relative to men [9].

This paper presents a global overview of the descriptive epidemiology of LC in relation to recent tobacco control policies, using the GLOBOCAN mortality estimates for the year 2020 provided by the International Agency for Research on Cancer (IARC) [10]. In addition, we provide projections of the future mortality burden according to different temporal scenarios to the year 2040, estimating the expected future LC deaths according to levels of Human Development Index (HDI).

## Data Sources and Methods

The number of deaths from, cancers of the lung (ICD-10 C33-34, including trachea and bronchus) were extracted from IARC’s GLOBOCAN 2020 database for 185 countries or territories, by sex and 18 age groups (0-4, 5-9, …, 80-84, 85 and over) [2,10,11]. Corresponding population data for 2020 were extracted from the United Nations (UN) website [12]. The data sources and hierarchy of methods used in compiling the cancer estimates have been described in detail elsewhere [10]. In brief, the GLOBOCAN estimates are assembled at the national level using the best available sources of cancer incidence and mortality data within a given country. The methods used to derive the 2020 estimates corresponding to those used to derived for previous years [13,14,15] where applicable, priority is given to short-term predictions and modelled mortality to incidence (M:I) ratios, while validity is dependent on the degree of representativeness and quality of the source information [10].

We present figures based on the estimated deaths in 2020, as well as two summary measures using direct standardization, namely the age-standardized mortality rate (ASR) per 100,000 person-years based on the 1966 Segi-Doll World standard population [16,17] and the cumulative risk of dying from cancer before the age of 75 expressed as a percentage, assuming the absence of competing causes of death [18]. These measures allow comparisons between populations adjusted for differences in age structures. We also provide a prediction of the future number of LC deaths worldwide for the year 2040, based on demographic projections and scenarios of uniformly increasing (+3%, +2%, +1%), stable (0%) or decreasing (−1%, -2%, -3%) rates annually from the baseline year of 2020. The possible impact of COVID-19 pandemic was not taken into consideration during the calculations.

The results are presented by country, and aggregated across 20 UN-defined world regions [12] and according to the UN’s four-tier HDI in 2020 [19], as a means to assess the cancer burden across four levels of development (low, medium, high and very high HDI). Throughout, we use the terms *transitioning, emerging* and *lower HDI* countries/economies as synonyms for nations classified as low or medium HDI, and *transitioned* or *higher HDI* countries/economies for those classified as high or very high HDI.

The Global Cancer Observatory (GCO, https://gco.iarc.fr) includes facilities for the tabulation and graphical visualization of the GLOBOCAN database, including explorations of the current [2] and future [20] burden for 36 cancer types.

Patient and Public Involvement: Patients or the public were not involved in the design, or conduct, or reporting, or dissemination plans of our research.

## Results

### Lung cancer mortality – national rankings 2020

Figure 1 presents global maps that indicate LC’s position in terms of deaths relative to other common tumours at the national level by sex for the year 2020. In 2020, LC ranks first in terms of cancer death in half, or 93 of the 185 countries included in GLOBOCAN, and either 2^nd^ or 3^rd^ in 37 countries in men. LC is a major contributor to cancer mortality around the world, including America, greater-Europe, Northern Africa, and across the Asian-Pacific region. There is a somewhat less dominant role at present in South America and Sub-Saharan Africa (but not South Africa). In women, the impact is lesser but still very much in evidence; the disease ranks as the leading form of cancer death in 25 countries including those within North America, Northern, Western and Southern-Central Europe, Eastern Asia and Australia/New Zealand. LC mortality ranks as the 2^nd^ or 3^rd^ leading form of cancer mortality in 54 countries worldwide in women.

**Figure 1:**
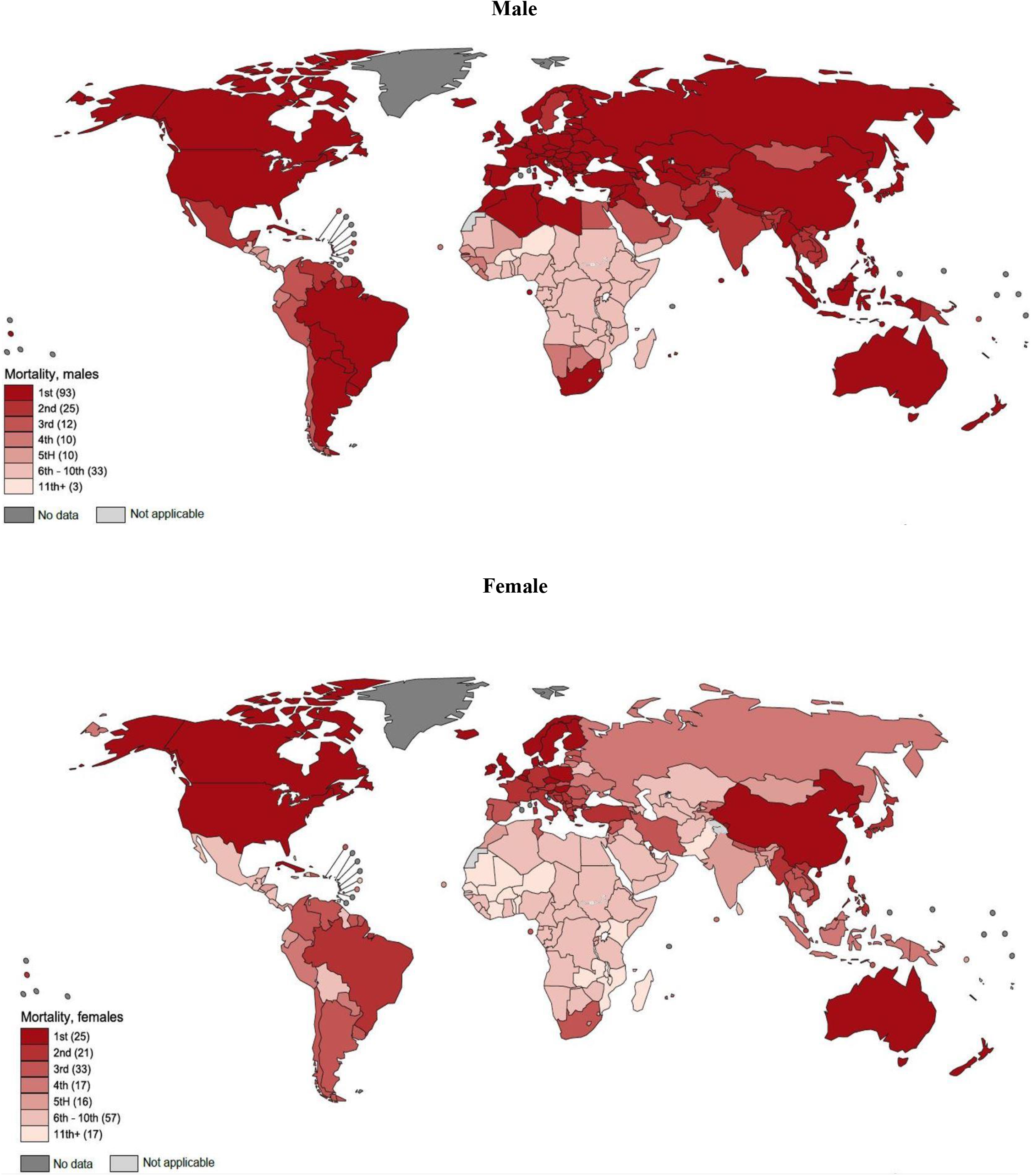
Lung cancer mortality compared with mortality from other causes of malignant neoplasms, 2020, Male-Female.

There is at least a 20-fold variation in mortality between the sexes, with rates uniformly higher among men (Figure 2). Male mortality rates are higher in Eastern and Southern Europe (especially in Hungary and Serbia with rates of 60 per 100,000), Eastern Asia (particularly the Democratic People’s Republic of Korea) and Polynesia and Micronesia, while rates are lower in Central America, South-Central Asia and most parts of sub-Saharan Africa. The highest female rates are observed in Northern America, Northern and Western Europe, and Australia/New Zealand specifically in Canada, Denmark and the Netherlands, respectively. Relatively low rates are observed in Western-, South-Eastern Asia and across the African continent, other than for South Africa.

**Figure 2:**
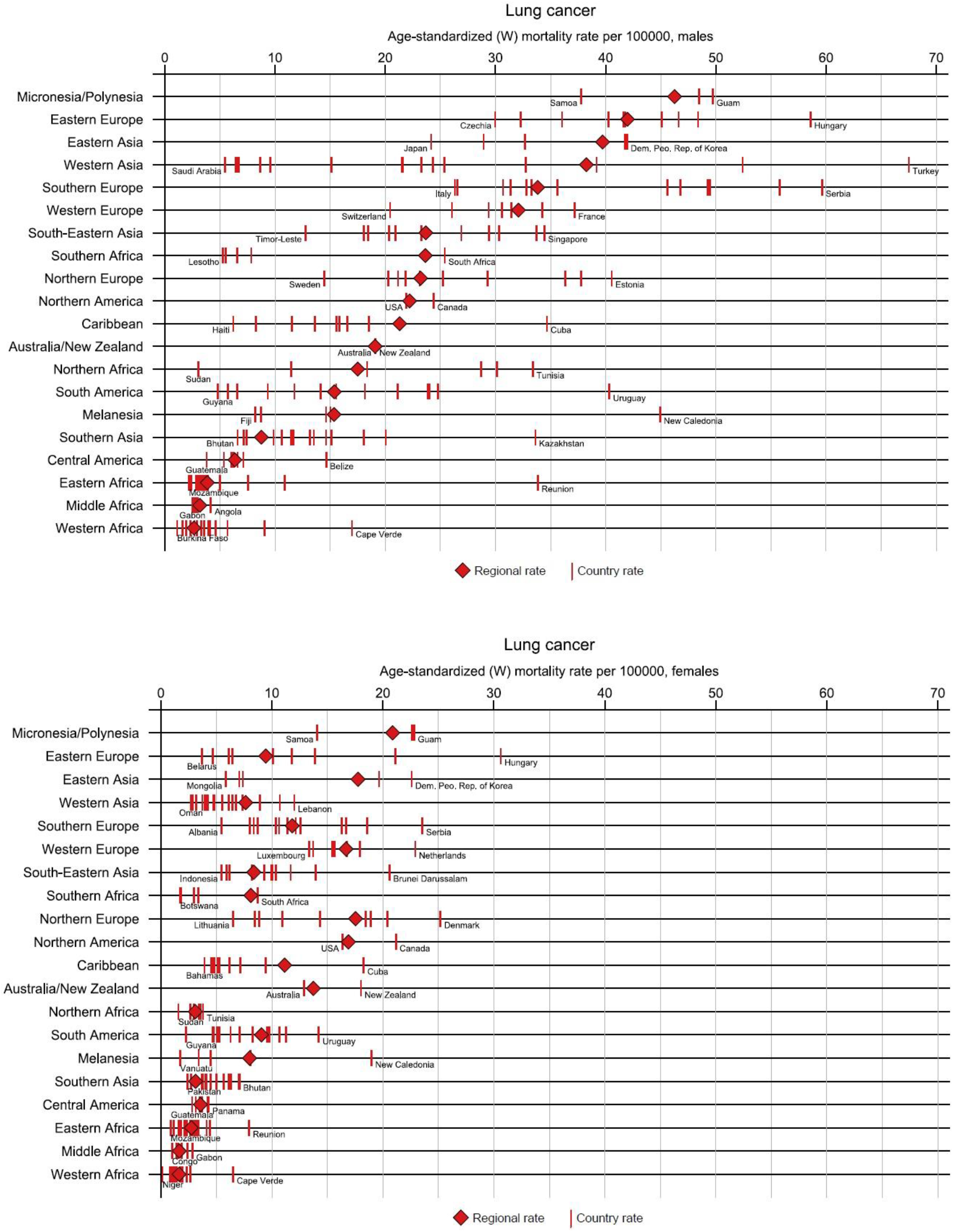
Lung cancer age-standardized mortality rates per 100,000 by world regions and sex in 2020, Male-Female.

### Lung cancer mortality burden by 2040

If the current rates were to remain constant over the next two decades, LC will claim around 2 million male deaths in 2040, compared to 1.2 million in 2020 (Figure 3). For women, the corresponding deaths are approximately half of their male counterparts: a predicted increase to 1 million in 2040 from 600 000 deaths in 2020. The projection also shows the different scenarios considering the changing rates per year between -3% and +3% based on plausible scenarios of the smoking epidemic in the short-term future; global declines in the number of LC among males but increases for female are perhaps the more realistic scenarios, with national or regional exceptions. Taking this trends-based prediction into account, the predicted number of deaths due to LC for men will likely range between 1.1 and 1.6 million and for women between 1.2 and 1.8 million by 2040. Deaths will markedly increase for both sexes in countries with the lowest HDI, even in the best-case trend scenario (Appendix Figure 1a-h).

**Figure 3:**
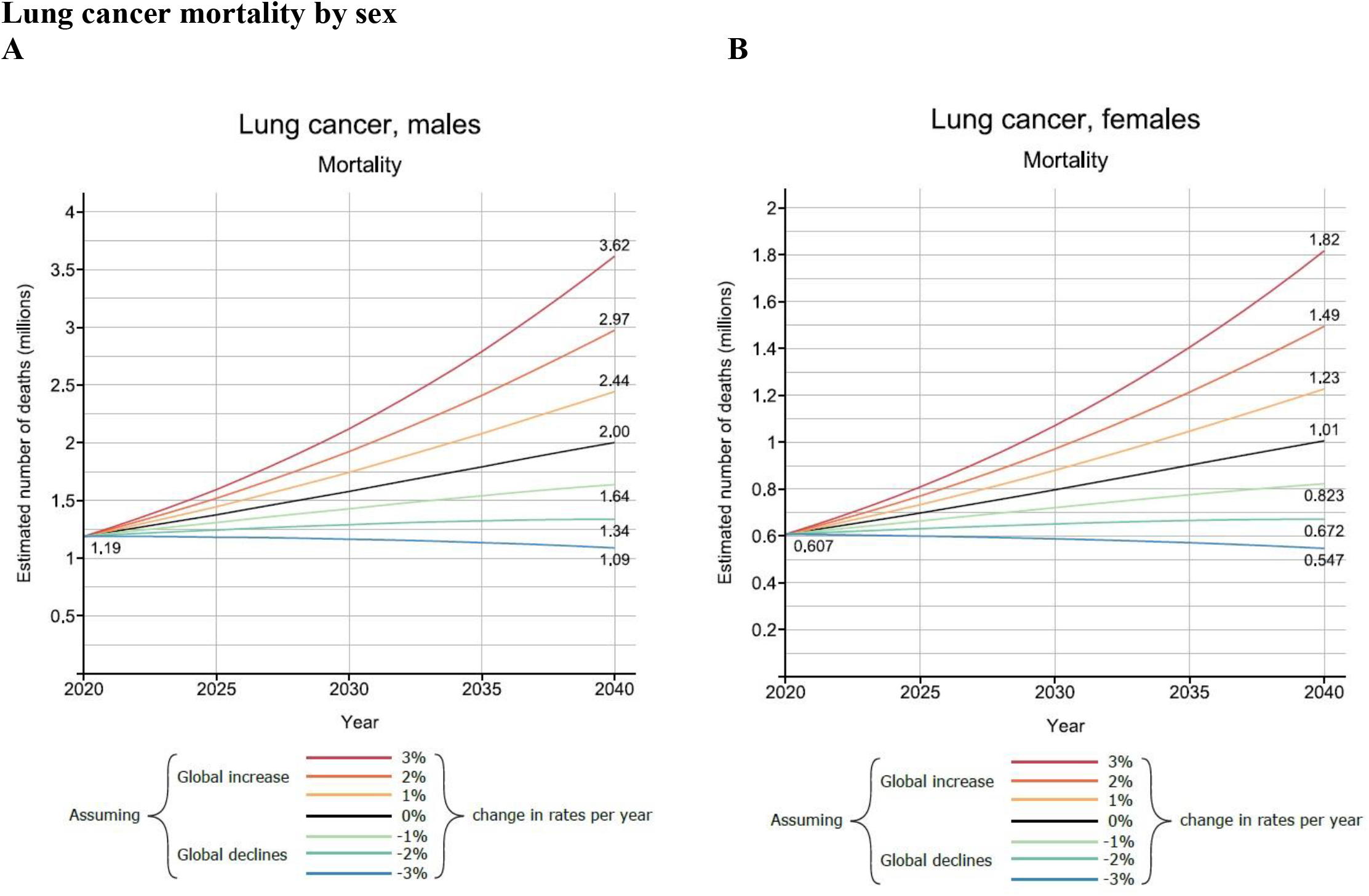
Lung cancer mortality projections worldwide from 2020 to 2040 by sex and the Human Development Index (HDI)

## Discussion

This study highlights the present geographic diversity in LC mortality worldwide, by sex and by level of human development. Countries with low HDI tend to have low LC mortality rates but may anticipate a higher mortality burden by 2040. For higher HDI countries, the burden of the disease is higher among men, but future trends suggest an increasingly greater proportion of the cancer burden will be seen among for females. These different scenarios are due to the impact of historic smoking trends and the increasingly widespread application of tobacco control measures in the last decades [21]. While there is an expectation that LC mortality will increase in transitioning countries given there is less implementation of effective tobacco control, there is a positivity in the findings of the Global Tobacco Control Report: the number of people now living in countries with at least two anti-tobacco policies in place rose from 3.5 billion in 2018 to 4.4 billion in 2020 – up from 45% of the world’s population to 56% in two years [22].

Past smoking histories of nations are a key determinant of the current magnitude of LC in many populations worldwide, as described by the classical model of the global smoking epidemic, first introduced by Lopez et al [4]. In the model, the effect of different smoking patterns was captured by four stages in the population, by an earlier adoption of the habit in men compared with women, and by the progressive adoption among lower socioeconomic classes, where the habit continues to be an underlying cause of the marked inequalities seen in different educational groups [23]. Lopez et al. initially applied the hypothesis on just a few developed countries [24], but later was tested on greater geographic scales [25,26]. Nevertheless, as smoking prevalence and subsequent LC rates began to peak and decline among men in many populations over the last decades, a key focus has been the deteriorating public health situation affecting women, where in many settings, rates of LC mortality have continued to rise. This raises several relevant biological, epidemiological and sociological concerns [27], including: the changing distribution of the main histological subtypes of LC over time [28], the extent to which females adopted the habit of smoking and their vulnerability to the tobacco industry [29,30], the impact of such a transition in diminishing gender differences in disease burden worldwide [31] and the effects of different political systems on the health awareness of individuals [32,33]. The impact of these factors is reflected in comparisons of between-country LC mortality rates; for example, the current rate differences in Eastern vs. Northern European countries.

Smoking is of course not the only risk factor for LC. There is strong evidence of a relation with other factors, including air pollution, climate change [34] and other occupational risk factors such as asbestosis and indoor exposure to cooking fumes etc [35]. The highest exposure to ambient air pollution is the characteristic of mainly countries in transition, where only modest reductions in burden will occur in the most polluted countries unless fine particulate matter (PM 2.5) values are decreased substantially [36].

Several other studies have aimed to forecast the future lung cancer burden in very high HDI countries e.g., the US [37] and the UK [38] with contradictory findings. While the steeply declining mortality in the US for both sexes until 2040 fits within the framework of the global smoking epidemic, the rising deaths reported in the UK for men and women until 2035 somewhat contradict previous findings. One explanation could be the rapidly ageing population age structure, which can increase the number of these non-standardized figures. Alternatively, these projections do not take into account the changing smoking prevalence in the past as a key determinant of present and future lung cancers. Our GLOBOCAN 2020 forecasts do not consider these either, however, provide possible scenarios on the basis of uniform increases or decreases in rates may help provide a realistic overview of the changing future burden of LC.

Another limitation of this study is the large variability in the availability and quality of cancer mortality data. Most African and some Asian countries suffer from weak mortality statistics systems. In GLOBOCAN, in countries where mortality series were not available from national vital registration sources, the predominant means of the estimation of rates were from corresponding national incidence estimates via modelling, using incidence-to-mortality ratios derived from cancer registries in neighbouring countries.

With over three million deaths predicted by 2040 in the absence of additional interventions according to the finding of this study, it is imperative to emphasize primary prevention as the most cost-effective strategy of tobacco control. It has been shown that raising the price of cigarettes through increased excise taxes can bring marked reductions in cigarette consumption [39]. Besides this, developing adaptive tobacco control strategies that target different subgroups is imperative; anti-tobacco strategies should urgently target women in settings such as the EU, in order to halt their rapidly increasing risk of LC, and prevent unnecessary, premature deaths among future generations of women [40]. In Sweden, as an example, policies such as those directed at health promotion have been implemented in a gender-specific way with a focus on young and pregnant women. Scotland also has gender-specific programs, such as the Women, Low Income, and Smoking Project [41]. Amos and Haglund (2000) have emphasized that building support for female-centered tobacco control programs through partnerships will be vital to achieve success [30]. Furthermore, Amos (1996) and Mackay and Amos (2003) draw attention to the situation of women in transitioning countries with presently low levels of cigarette smoking among women [29]. In these countries, smoking among girls is already on the rise, women’s spending power is increasing, cigarettes are becoming affordable, and women are more exposed to the marketing strategies of tobacco companies, in an environment where cultural constraints are weakening and female-specific quitting programs are rare [8].

A package of measures to suppress tobacco consumption in a given population has been recommended through continued efforts to increase the proportion of ex-smokers, with a focus on younger generations [42]. This could perhaps be achieved by implementing coordinated smoking prevention and control strategies from an early age, in the form of educational programs in schools. Other measures that could be introduced include community intervention programs, mass media campaigns and further legislation to ban smoking in public places. One of the main problems is that young people react very differently to anti-smoking messages compared to adult long-term smokers [42]. The harm-reducing role of e-cigarettes and aid to smoking cessation has been proposed [43], however their impact on future LC mortality is not yet known [44]. Successful programs have also been implemented in rapidly emerging economies such as Brazil, where a reduction in smoking prevalence were observed after the ratification of the WHO Framework Convention on Tobacco Control (FCTC) in 2005, and the adoption of a national ban on tobacco advertising, a national comprehensive smoke-free policy, large pictorial health warnings on cigarette packages, and continuous increases in taxes and prices of tobacco products [45]. Other factors may influence the future burden of LC such as the potential introduction of screening in high-risk populations. In a recent trial, LC mortality was significantly lower among those who underwent volume computed tomography (CT) screening than those who did not participate [46]. Screened patients benefitted from a substantial shift to lower-stage cancers at the time of diagnosis as well as more frequent eligibility for curative treatment (mainly surgery) [47]. However, concerns have been raised about the potential for overdiagnosis in lung-cancer screening.

In summary, this paper has identified marked geographic variations in the current LC burden worldwide and provided potential scenarios regarding the short-term future LC deaths up until 2040. Gredner et al., have illustrated the great potential of comprehensive implementation of tobacco control policies in Greater-Europe, with over 1.6 million LC cases preventable over a 20-year period through the highest-level implementation of tobacco control policies [48]. There is therefore much we can do to halt the rising deaths from LC – as well as many other forms of cancer and non-communicable diseases –through the successful implementation of tobacco control policies.

## Disclosure

Where authors are identified as personnel of the International Agency for Research on Cancer/World Health Organization, the authors alone are responsible for the views expressed in this article, and they do not necessarily represent the decisions, policy, or views of the International Agency for Research on Cancer/World Health Organization.

## Data Availability

All data produced are available online at the webpage of the Global Cancer Observatory.

https://gco.iarc.fr/

## Role of the founding source

PN: This study was supported by the Topic Excellence Program (TKP2020-NKA-26, TKP2021-EGA-44), the National Laboratories Program (National Tumor Biology Laboratory-2022-2.1.1-NL-2022-00010), and Tasks Related to the National Public Health Strategy (IV/4925/2021/ EKF).

## Authors’ contribution

AW: literature search, data analysis, writing – original draft; EM: writing – review & editing; JV: methodology, data collection, figures, visualisation; ML: visualisation; MP: writing – review & editing; HR: writing – review & editing; DS: writing – review & editing; PN: writing – review & editing, funding acquisition; IK: writing – review & editing; IS: writing – review & editing; FB: methodology, conceptualisation, data analysis, writing – original draft

## Data sharing

The data that support the findings of this study are available at the Global Cancer Observatory (GLOBOCAN) at https://gco.iarc.fr/

Data are available in a public, open access repository.

## Ethics Approval Statement

This study does not involve human participants and animal subjects.

