## Supplemental Table 1 for "Lung cancer mortality in the wake of the changing smoking epidemic: a descriptive study of the global burden in 2020 and 2040"

Supplemental material - Appendix

**Figure 1: Lung cancer mortality compared with mortality from other causes of malignant neoplasms, 2020, Male - Female**

The boundaries and names shown and the designations used on this map do not imply the expression of any opinion whatsoever on the part of the World Health Organization concerning the legal status of any country, territory, city or area or of its authorities, or concerning the delimitation of its frontiers or boundaries. Dotted and dashed lines on maps represent approximate border lines for which there may not yet be full agreement.

Data source: Globocan 2020, Map production: CSU, World Health Organization

Ferlay J, Ervik M, Lam F, Colombet M, Mery L, Piñeros M, Znaor A, Soerjomataram I, Bray F (2020). Global Cancer Observatory: Cancer Today. Lyon, France: International Agency for Research on Cancer. Available from: https://gco.iarc.fr/today, accessed [13 April 2022].

### Appendices

**Lung cancer mortality by sex - very high HDI countries**

**A**

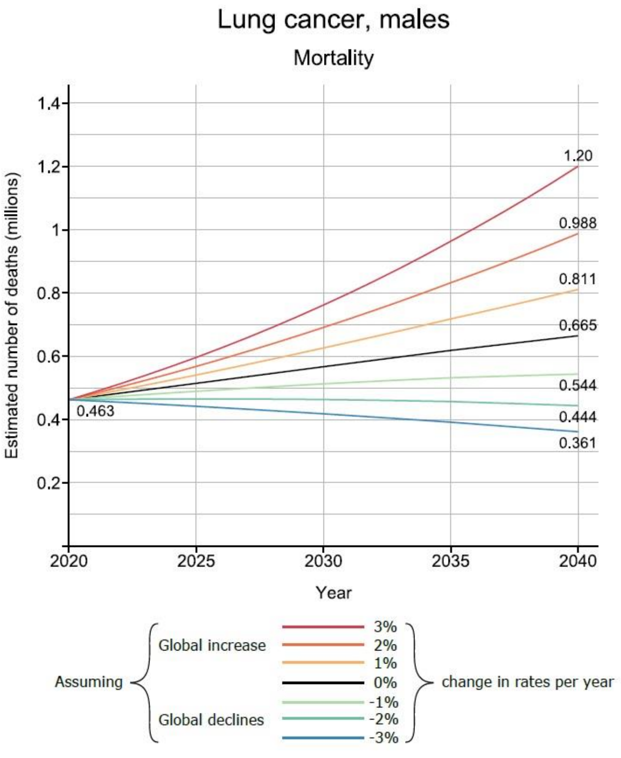


**B**

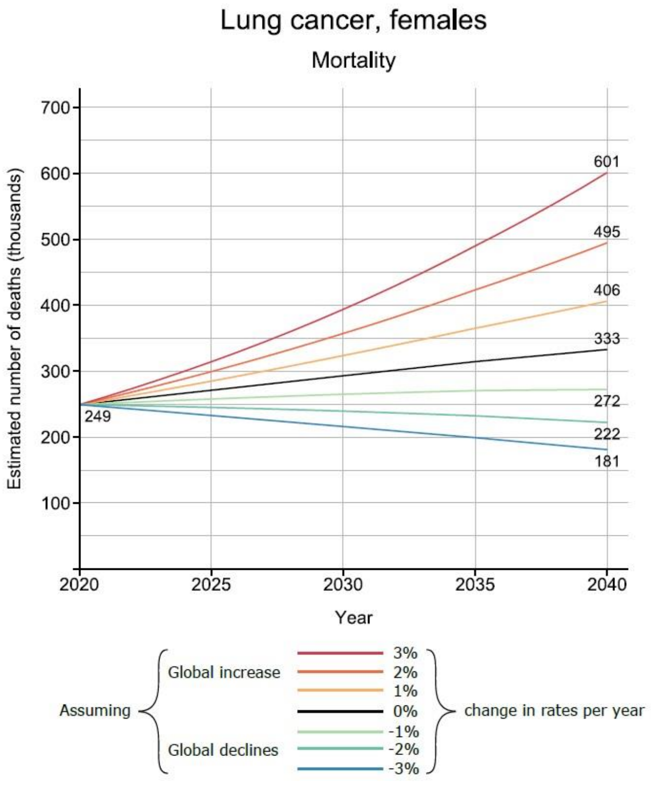

### Lung cancer mortality by sex - high HDI countries

**C**

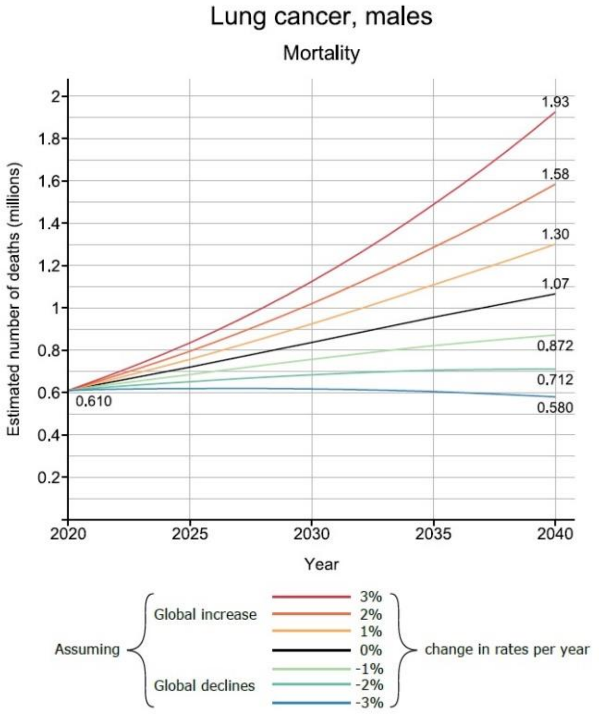


**D**

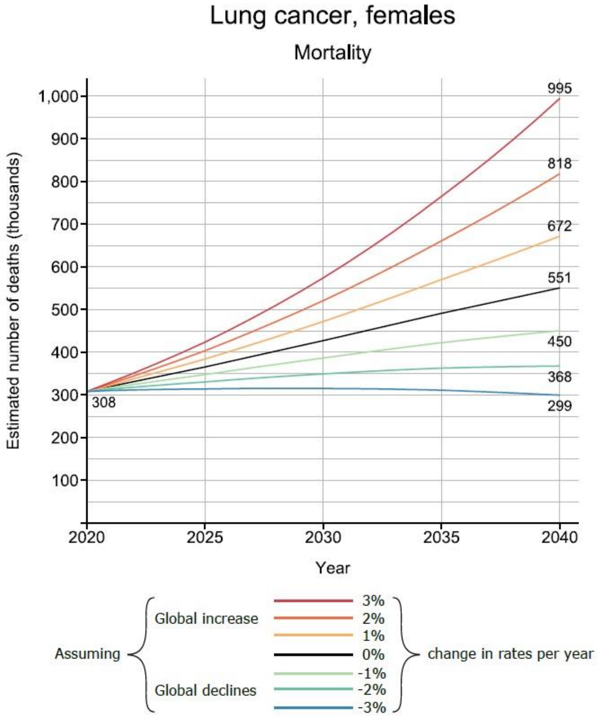


### Lung cancer mortality by sex - medium HDI countries

**E**

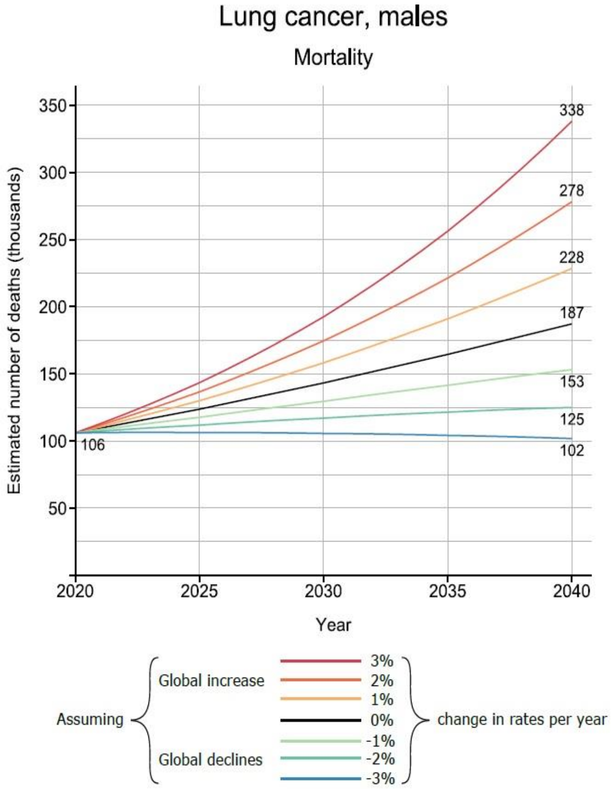


**F**

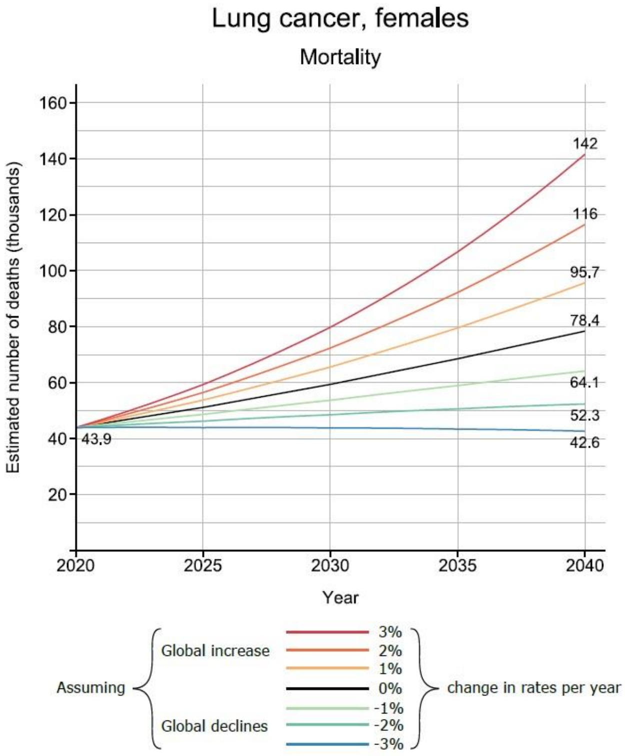

**Lung cancer mortality by sex - low HDI countries**

**G**

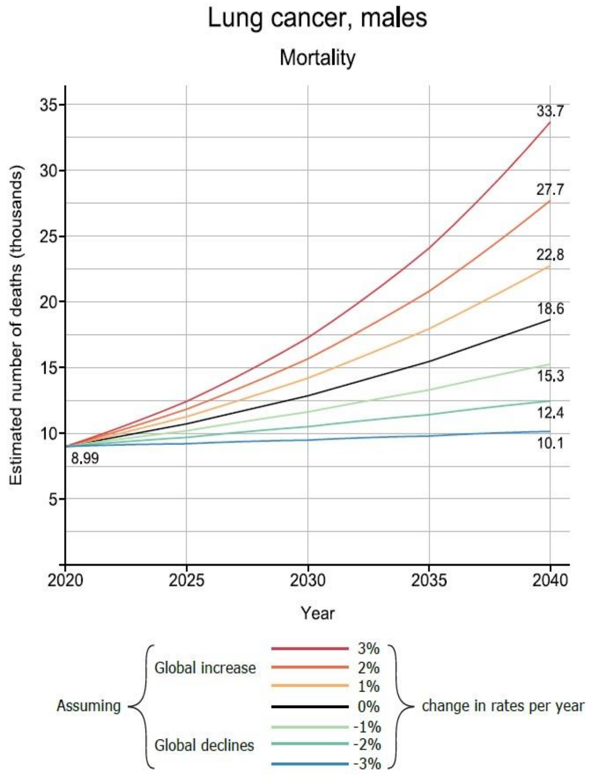


**H**

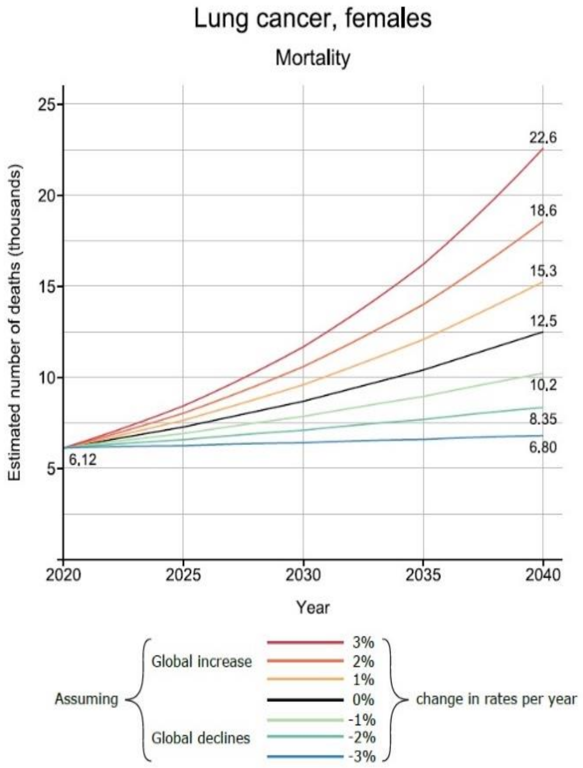
